# Spatial and temporal analysis of cholera cases in high-risk areas of Kenya

**DOI:** 10.1101/2023.10.13.23296989

**Authors:** Lydia M Mageto, Nyamai Mutono, Gabriel Aboge, Peter Gathura, Emmanuel Okunga, Annastacia Muange, Cecilia Mbae, SM Thumbi, Samuel Kariuki

## Abstract

Cholera outbreaks remain an important public health challenge in Kenya, especially in areas with poor sanitation and hygiene including urban informal settlements, refugee camps and rural areas bordering large water bodies. Even in endemic settings, the distribution of cases exhibits spatial and temporal variations. Utilizing a Poisson discrete space-time scan statistic (SaTScan), this study investigated the temporal trends and the nature of spatial spread of cholera within selected high-risk areas in Kenya. The study was conducted in an urban informal settlement in Nairobi (Mukuru), a refugee camp in Northern Kenya (Dadaab) and the four counties bordering Lake Victoria region. Retrospective cholera line list data from January 2012 to December 2022 in the selected high-risk areas was used. SaTScan v10.1.2 was used to carry out Spatiotemporal analysis and generate spatial clusters. Throughout the study period, a total of 7,372 cholera cases were reported, corresponding to a lower annual incidence rate of 12.2 per 100,000 people compared to a mean annual incidence of 25 cases per 100,000 population previously reported in Kenya. The highest number of cases (n=5934) were reported between 2015 and 2018 with an annual incidence rate of 27.0 per 100,000 people, indicating a relative risk (RR) of 7.22 and a log-likelihood ratio (LLR) of 3015.17 (p< 0.001). The risky clusters (RR>1) were in Dadaab, Fafi, Suna West, Nyatike, Ugunja, Ndhiwa and Suna East sub counties with annual cases of 111.6, 164.1, 28.4, 19.9, 19.4, 14.0 and 13.9 per 100,000, respectively. The sub-counties of Nyakach, Nyando, Rachuonyo East, Kisumu East and Kisumu Central were reported as low-risk clusters, with a relative risk of 0.055 and an annual incidence rate of 1.1 cases per 100,000 individuals. Out of the thirty-two sub-counties included in the study, ten of them did not report any cases of cholera during the study period. Cholera cases waxed every three years in the selected high-risk areas. This data on hotspots specific to endemic settings forms a basis for prompt public health response and resource allocation by prioritizing the significantly high-risk clusters to control and eventually eliminate the disease.

## Introduction

Cholera caused by *Vibrio cholerae* (*V. cholerae*) has been incriminated as one of the major causes of mortality predominantly in areas of poor sanitation and hygiene globally. In 2015, the global burden of cholera was estimated at 1.3 – 4 million infections and 143,000 mortalities. The disease is endemic in 69 countries with 60% and 29% of the population residing in Africa and Asia respectively. The bacterial pathogen is spread through fecal-oral route either directly from person-to-person or indirectly through consumption of contaminated food or water (Sack et al., 2003). *A*ccording to World health organization (WHO), about 1 in 10 people who contract cholera develop severe clinical signs in 12 hours to 5 days characterized by vomiting, watery diarrhoea, leg cramps, restlessness, and irritability (WHO, 2017). WHO recommends intravenous or oral hydration as the main treatment for cholera in case of mild to moderate cases, while for severe cases and in patients with underlying conditions, prompt antibiotic treatment using drugs such as doxycycline is recommended. In case treatment is not promptly initiated, death may occur within hours after onset of symptoms (Adagbada et al., 2012).

Since the resurgence of cholera in 1970, 50% of the global cholera cases have been reported in Sub-Saharan Africa from 1980 to 2011 (Mengel et al., 2014). In Kenya, the disease has been widely reported primarily affecting the poor and vulnerable communities living in informal settlements in peri-urban areas and refugee camps where overcrowding, inadequate sanitation and water shortages are frequent (Stoltzfus et al., 2014; Hounmanou et al., 2016). A recent cholera hotspot mapping study done in Kenya identified urban informal settlements, large refugee camps, pastoral areas, arid and semiarid lands, areas bordering the lake region and Mwea irrigation scheme as high priority (Kiama et al., 2023)

Inadequate hygiene practices, lack of clean water, insufficient toilet facilities and ineffective waste treatment significantly increase the risk of cholera outbreaks in informal settlements. Additionally, improper solid waste management aggravates the problem by causing clogged drainage systems and flooding, further increasing the risk of cholera outbreaks. Overcrowding is a major issue in refugee camps. Refugees reside in small tents and makeshift homes with limited access to proper sanitation and hygiene facilities. These conditions significantly increase their vulnerability to cholera infection (International Rescue Committee, 2017). For instance, Individual and communal taps are the primary water sources in the camps. However, use of unclean water storage containers increases the risk of infection. In addition, poor handwashing facilities and improper human waste disposal practices have previously been associated with a heightened risk of cholera outbreaks in such settings. Even though Pit latrines are used for human waste disposal, they are often inadequate (Luby et al., 2020).

Fishing is the main economic activity for people living in the Lake region. Individuals engaged in subsistence fishing often migrate and settle temporarily in various regions. Limited access to clean water and inadequate hygiene and sanitation practices are major risk factors for cholera outbreaks in these temporary settlements (Mills et al., 2011; Bene & Neiland, 2003) . Increased cholera outbreaks in the lake region have also been associated with consumption of lake water (Bwire et al., 2017; Bompangue et al., 2008). Additionally, during the rainy season, environmental *V. cholerae* attach themselves to plant surfaces, algae, and zooplankton, acting as reservoirs and sources of disease transmission (Khonje et al., 2012; Rebaudet et al., 2013).

Since 1971 when the first outbreak was reported in Kenya, a number of significant cholera outbreaks have been reported (Kigen et al., 2020;Mutonga et al., 2013; Scrascia et al., 2009; Shikanga et al., 2009; Mugoya et al., 2008 ; Shapiro et al., 1999; Tauxe et al., 1995). Recent estimates show that up to 4.9 million people live in cholera prone areas in Kenya commonly known as hot spots (Kiama et al., 2023). The 2022 outbreak in Kenya was reported in 25 (53%) counties according to the Cholera outbreak situational reports by Kenya’s Ministry of Health (https://reliefweb.int/report/kenya/kenya-cholera-outbreak-operational-update-appeal-no-mdrke054-22-july-2023).

Globally, there have been efforts to reduce cholera mortalities by 90% and eventually eradicate the disease in 20 out of the 47 high burden countries by 2030 (Global Task Force on Cholera Control, 2017). Kenya is among the 20 countries targeted for cholera eradication. The Global Task Force on Cholera Control, 2017 outlines a roadmap towards cholera eradication by 2030 through improved Water, Sanitation and Hygiene (WASH), use of Oral Cholera Vaccines (OCVs), hastening disease surveillance in high-risk areas, and early detection and prompt response to outbreaks. Kenya’s strategy for cholera control is to deploy OCVs in addition to improved WASH facilities to populations living in cholera hotspots, areas that carry the highest disease burden and majorly contribute to disease spread. With such targeted efforts to specific areas, the interventions will reach communities that need them most(Golicha et al., 2018).

Since then, significant progress has been made, as Kenya has undertaken cholera hotspot mapping using Water, Sanitation, and Hygiene (WASH) measures, along with other epidemiological indicators, as an integral component of the country’s new cholera elimination plan for the period 2022-2030(Kiama et al., 2023). Attempts have been made to investigate spatio-temporal trends of cholera across the world (Domman et al., 2017). A few countries in Africa including Zimbabwe and Ghana have investigated the temporal trends and the nature of spatial interaction of cholera (Ngwa et al., 2016; Osei & Stein, 2018; Luque Fernandez et al., 2012). Furthermore, studies on spatio-temporal trend of cholera have been extensively done in Cameroon, Uganda, Zambia, and Tanzania to identify cholera clusters and provide valuable insights for implementing intervention measures (Mwaba et al., 2020; Hounmanou et al., 2019; Bwire et al., 2017; Ngwa et al., 2016).

In Kenya, the cholera hotspot mapping study identified urban informal settlements, refugee camps, and areas bordering large water bodies as high-risk areas for cholera outbreaks. However, there is still insufficient data regarding the spatial and temporal clustering of the disease specific to high-risk sub counties (Kiama et al., 2023). This has hampered efforts to implement effective intervention measures for cholera prevention and control in hotspots.

Therefore, the goal of this study was to identify spatial and space-time clusters of cholera in specific high-risk areas of Kenya by employing the spatial scan statistic, SaTScan. The findings from this study will provide the government with data that will assist targeted intervention and provide an early warning for more effective response measures. It will also inform public health intervention towards control and eventual elimination of cholera in Kenya. This will enable the country to meet its Sustainable Development Goal of good health and wellbeing by 2030.

## Methods

### Study area

The study was conducted in three areas including i) Dadaab refugee camp, ii) Counties bordering Lake Victoria region (Kisumu, Siaya, Migori and Homabay) and iii) Mukuru informal settlement in Nairobi. The Lake Victoria region of Kenya and Mukuru informal settlements mostly receive long rains between March and May and short rains in October to December while Dadaab is semi-arid throughout the year.

Mukuru informal settlement has a population of 242,941 people with 97,890 households with an average of 466 people per acre while the Lake Victoria region has a human population of 4,397,143 (Kenya National Bureau of Statistics, 2019). Dadaab refugee camp, situated in Garissa County, is comprised of three camps (Dagahaley and Ifo in Dadaab sub county, and Hagadera in Fafi sub county). The camp has a population of approximately 240,984 people, primarily refugees from Somalia (UNHCR operational portal, 2023). These areas were purposively selected because of their increased risk to cholera outbreaks due to overcrowding, inadequate access to clean water as well as poor sanitation and hygiene (Kiama et al., 2023; Mills et al., 2011; Bene & Neiland, 2003). A map of the study area is shown in Figure 1.

**Figure 1:**
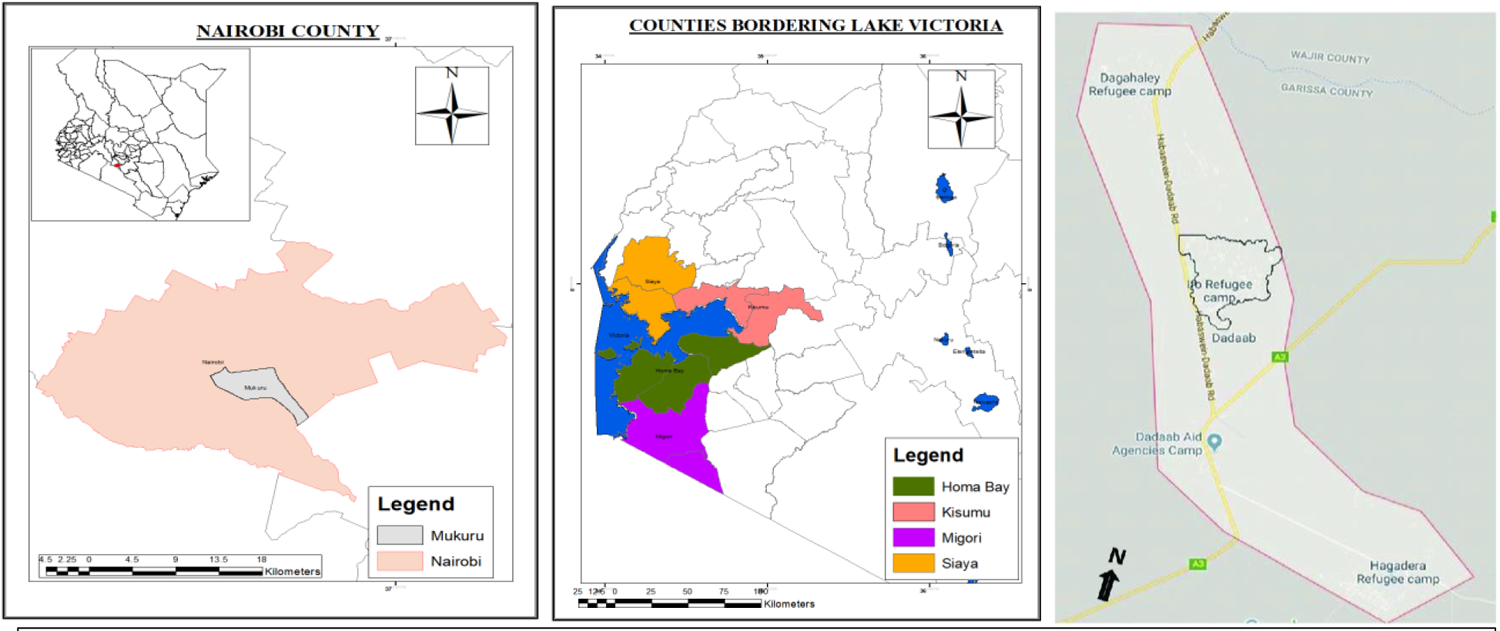
Map of the study areas in Kenya: Mukuru informal settlement, Counties bordering Lake Victoria region and Dadaab refugee camp.

### Data collection

Cholera data from 2012 to 2022 was acquired from the Division of Disease Surveillance and Response (DDSR) at Kenya’s Ministry of Health, at a sub county level. Cholera cases were either clinically or laboratory confirmed with the former based on symptoms which included watery diarrhea occurring at least three times a day, with or without vomiting. Laboratory confirmed cases were identified by isolation of Vibrio cholerae O1 or O139 in the stool. Both suspected and confirmed cases were included in the study. Population data at sub county level was extrapolated from the 2019 Kenya Population and Housing Census report (Kenya national bureau of statistics, 2019). The shape files of Kenyan Sub counties were obtained from an open data source. (https://geodata.ucdavis.edu/gadm/gadm4.1/shp/gadm41_KEN_shp.zip). Given the difference in size of sub county polygons, sub county Centroids were obtained using R version 4.1.0. The obtained data on sub county centroids, number of cholera cases and population per sub county was securely saved as Comma Delimited (CSV) files.

#### Organization of the dataset

Data containing the county, sub-county, total population in each sub-county, and the centroids of the sub-counties in the identified high-risk areas was recorded in Microsoft Excel. This data saved as CSV file, was then imported into SaTScan v10.1.2 software for conducting spatio-temporal analysis (SaTScan, 2005). The software required three specific data files to perform the analysis including: (1) Case file which contained data on both suspected and confirmed cholera cases from 2012 to 2022 (2) Population file with information on the number of people at risk in each sub-county obtained from the 2019 Kenya Population and Housing Census report (Kenya National Bureau of Statistics, 2019). (3) Coordinates file which had information about the specific geographical location of each sub-county. The analysis in this study was conducted at the sub-county level, and therefore the centroids of the sub-counties were used as the coordinates for each sub-county.

### Data analysis

#### Spatial scan statistical analysis

SaTScan version 10.1.2 was employed to detect and assess purely spatial, spatial variation in temporal trends or space-time disease clusters and test their statistical significance. SaTScan employed a Poisson-based model to calculate high risk clusters based on a Monte Carlo simulation (Block, 2007; Kulldorff, 2006).

### Purely spatial analysis

To identify purely spatial clusters, SaTScan imposed a circular window placed over the study region, centered on the centroid of each sub county, and moved across the region. The size of the window was set at 50% of the total population at risk. Within each window, potential clusters were tested when the window was centered on the centroid of each sub county. When a new case was encountered within the window, the software calculated a likelihood function to assess the elevated risk within compared to areas outside the window based on a protocol by Kulldorff, 2006.

The likelihood function was maximized over all windows with an aim of identifying the window that formed the most probable cluster and was least likely to have occurred by chance. The likelihood ratio for the window was recorded as the Maximum Likelihood Ratio (MLR) test statistic. MLR identified the most likely clusters with a higher number of observed than expected cases. Distribution of the MLR under the null hypothesis as well as its corresponding P value were determined by employing a Monte Carlo simulation approach (Block, 2007).

### The space–time scan statistic

SaTScan used a cylindrical window with a circular geographic base to detect space-time clusters. This window was imposed by centering it around one of several possible centroids located throughout the study region. The radius of the window was continuously adjusted in size, including both the geographical area and the time interval. The window was systematically moved across space and time, exploring every possible geographic location and size, as well as considering each potential time interval. This process ensured that all combinations of space and time were examined to detect clusters (Kulldorff et al., 1998). In order to determine the significance of the space-time clusters detected, a likelihood ratio test statistic was constructed in a similar manner as for the purely spatial scan statistic. It compared the observed number of cases within the window to the expected number of cases under the null hypothesis, using a likelihood ratio approach (Kulldorff, 2001).

In this study, a spatial window with a maximum radius of 50% of the population at risk and a maximum temporal window of 50% of the study period was employed. This approach allowed for the detection of both purely spatial clusters and clusters that exhibited spatial and temporal patterns. Additionally, the study conducted scans for the most likely clusters of various time lengths. These scans utilized a circular spatial window with a radius equivalent to 50% of the population at risk, along with a maximum temporal window of 90%. This approach allowed for the identification of clusters exhibiting purely spatial patterns, purely temporal patterns, as well as combined spatial and temporal patterns. The analysis assumed that the risk ratio of cholera was consistent both within and outside the identified window. Multiple circular windows of varying sizes, representing potential clusters, were generated for analysis. The analysis was conducted using a Poisson based model, and the significance level was determined with a threshold of P < 0.05, indicating statistical significance.

### Spatial variation in temporal trends scan statistic

The scanning window was purely spatial in nature in this statistic. Analysis of the temporal trend of cholera cases was computed both inside and outside the scanning window for each sub county based on a null hypothesis that the trend is similar within and outside the window. A likelihood ratio was calculated based on this hypothesis with an annual increase or decrease in risk. The cluster for which within the window temporal trend was least likely to be the same as outside the window temporal trend was considered the most apparent cluster. A negative number characterized a decreasing trend in risk while an increasing trend was linked to a positive number in the output.

## Results

### Descriptive spatio-temporal analysis

A total of 7372 cholera cases were reported from 2012–2022 in Dadaab refugee camp, Mukuru informal settlement and the four counties bordering Lake Victoria region, affecting 22 (69%) of the 32 sub-counties in the study area. The largest number of cases affecting 21 sub counties (n=2784) were reported in 2015 while the lowest were in 2020 (n=7) whereby only 1 sub county was affected (Figure 2). The 22 sub-counties where cholera was reported accounted for 11.5% (5.49 million people) of the total Kenyan population. Throughout the study period, Fafi sub-county experienced the highest burden of cholera with 2420 reported cases, followed by Dadaab with 2274 cases, and Suna West with 402 cases. Figure 2 presents the yearly distribution of cholera cases at the sub-county level. Zero cases in some sub-counties indicate a lack of reporting rather than absence of an outbreak.

**Figure 2:**
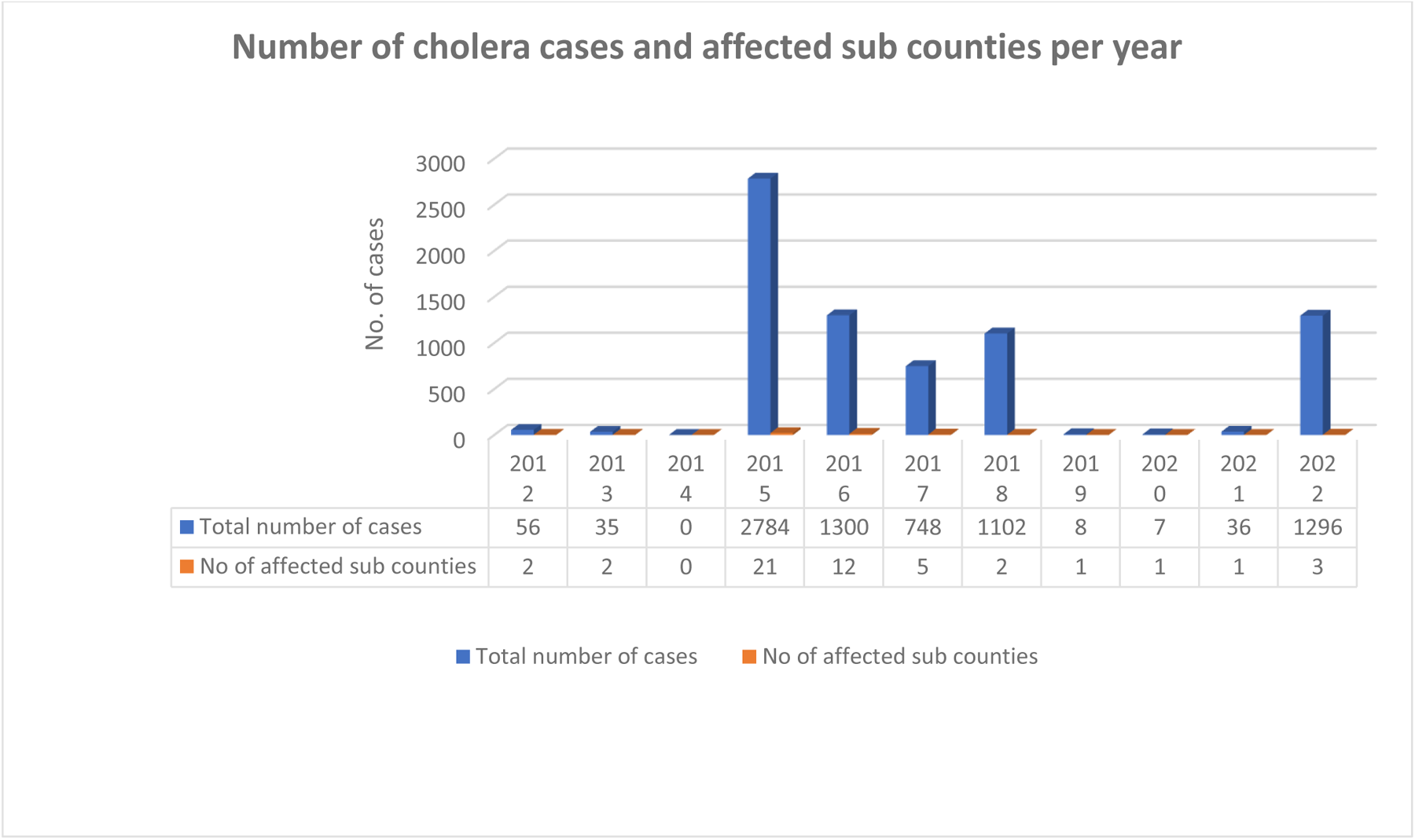
Distribution of cholera cases per year in selected high-risk areas in Kenya, 2012-2022

The analysis of temporal clusters with high concentration of observed cases identified a cluster interval for cholera cases between 2015 and 2018. The highest peak incidence rate was observed in 2015, with the largest difference between expected and observed number of cases. In contrast, the lowest number of cases was observed in 2020, falling below the expected number. These findings are graphically represented in Figure 3.

**Figure 3:**
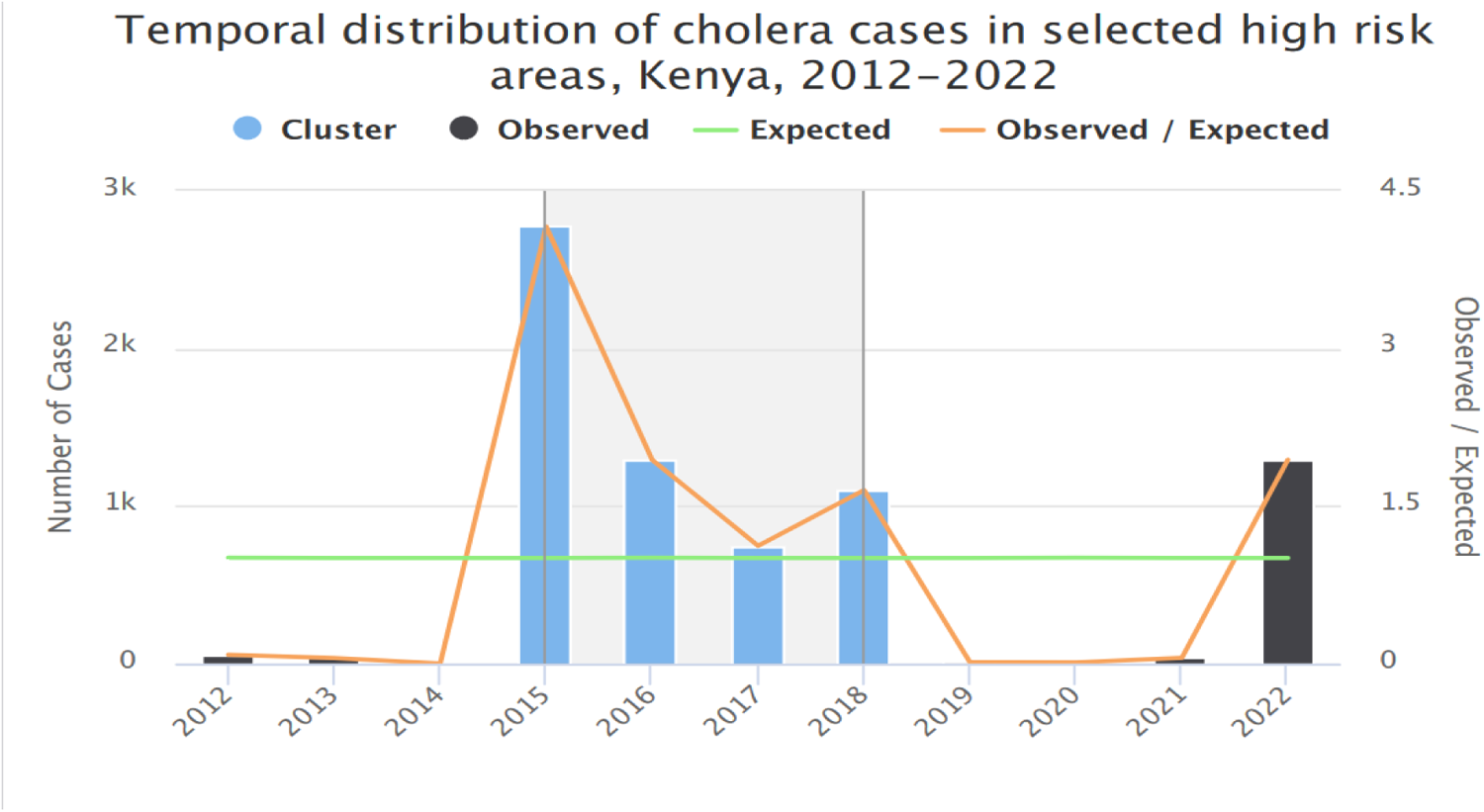
Temporal distribution of cholera clusters from selected high-risk areas, Kenya, 2012-2022

### Purely Spatial distribution of cholera in high-risk areas in Kenya

There were five significantly high-risk clusters with more observed than expected cases (Relative Risk > 1). These clusters were observed in Dadaab, Fafi, Suna West, Nyatike and Ugunja with annual cases per 100,000 people of 133.6, 23.5 and 19.4 respectively. People living in these sub counties were 1.60 to 28.37 times more likely to contract cholera compared to those residing elsewhere in the country. Approximately 728,585 people live in these high-risk sub counties. The risk of cholera infection was significantly lower in 16 sub counties namely Nyakach, Nyando, Rachuonyo East, Kisumu East and Kisumu Central as shown in Table 1 and Figure 4. The high and low risk clusters were significantly diverse among sub counties (p < 0.001).

**Table 1:** Purely Spatial clusters of cholera detected by retrospective spatial analysis using a Circular spatial window with a radius of 50% of the population at risk in high-risk areas, Kenya, 2012-2022.

| Name of subcounty | Population | Observed cases | Expected cases | Annual cases/100000 | Relative risk | Log Likelihood Ratio | P Value |
| --- | --- | --- | --- | --- | --- | --- | --- |
| Dadaab, Fafi | 319,292 | 4694 | 428.99 | 133.6 | 28.37 | 8679.669408 | 0.0001 |
| Suna West, Nyatike | 305,052 | 787 | 409.86 | 23.5 | 2.03 | 146.72 | 0.0001 |
| Ugunja | 104,241 | 222 | 140.05 | 19.4 | 1.60 | 20.78 | 0.0001 |
| Ndhiwa | 218,136 | 335 | 293.08 | 14.0 | 1.15 | 2.99 | 0.466 |
| Suna East | 122,674 | 188 | 164.82 | 13.9 | 1.14 | 1.60 | 0.925 |
| Nyakach,<br>Nyando,<br>Rachuonyo East,<br>Kisumu East,<br>Kisumu Central,<br>Muhoroni,<br>Rachuonyo<br>North, Kisumu<br>West, Suba<br>South, Rangwe,<br>Seme, Homabay<br>Town, Rongo,<br>Rarieda, Gem,<br>Awendo | 2,387,875 | 302 | 3208.26 | 1.1 | 0.055 | 3029.55 | 0.0001 |

**Figure 4:**
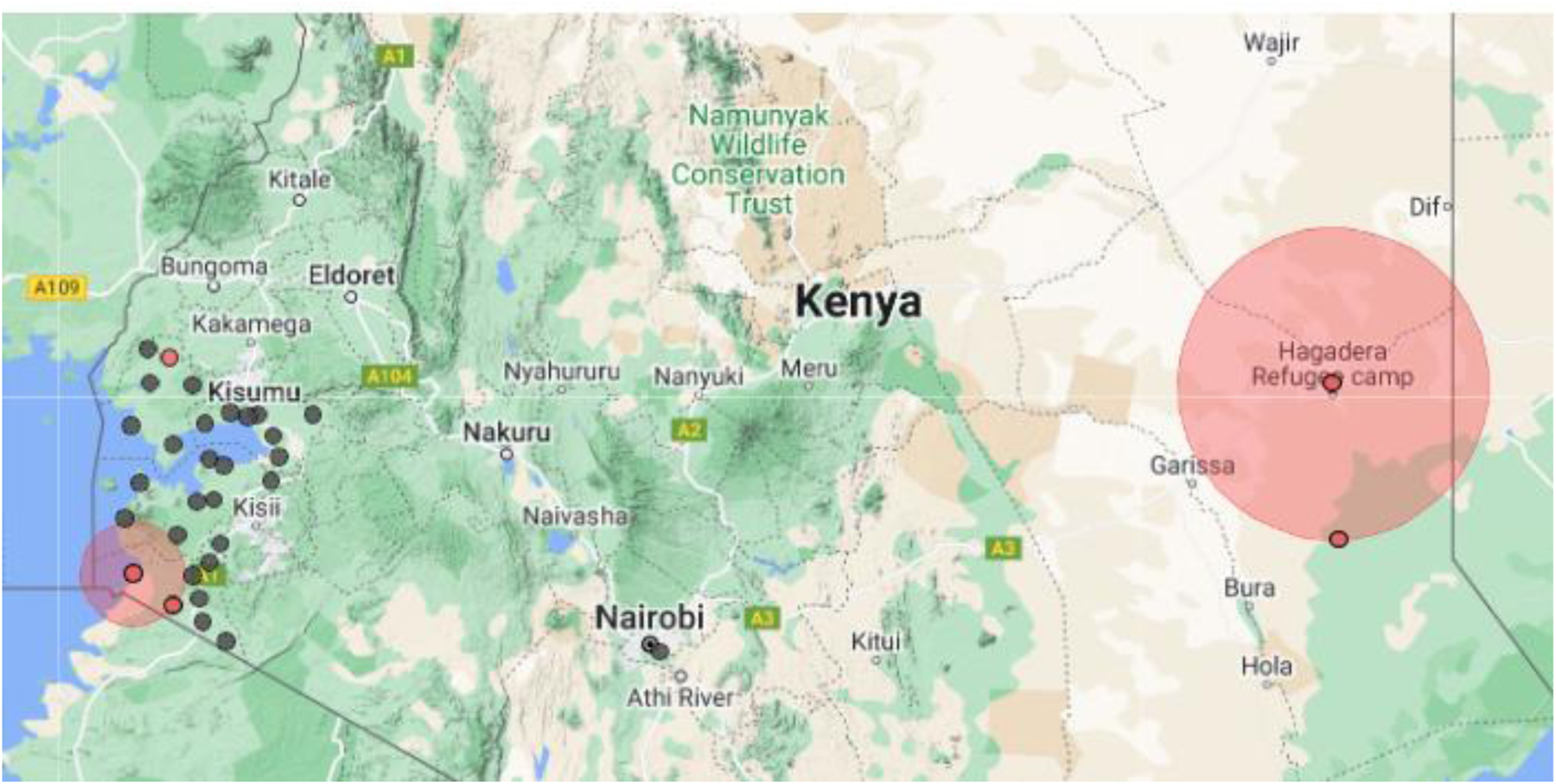
Map showing 3 clusters and 5 significant spatial cluster locations of cholera in selected high-risk areas, Kenya, 2012-2022

### Space–time variability of cholera

Space-time analysis was carried out to ascertain whether the high-risk clusters from purely spatial analysis were temporary or long term. The analysis was conducted using a circular spatial window with a radius of 50% of the population at risk. Additionally, a maximum temporal window of 50% was applied, which included purely spatial clusters. During the study period (2012 to 2022), Dadaab and Fafi sub-counties, located approximately 90 km apart, exhibited the highest number of annual cholera cases. The incidence rate in these sub-counties was 133.6 cases per 100,000 population, indicating a significant cholera burden. They also had the highest relative risk (RR=28.37, P<0.05). Dadaab and Fafi Sub counties remained statistically significant for high cluster of cholera cases throughout the year 2015-2018. One statistically significant secondary cluster encompassing Alego Usonga, Ugunja and Ugenya was detected in the year 2016. There was significant variation in detection of cholera cases annually in both time and space (p<0.001). This is shown in Table 2 and Figure 5 below.

**Table 2:** Significant high-risk spatial clusters of cholera in high-risk areas in Kenya, detected by retrospective space-time analysis using a Circular spatial window with a radius of 50% of the population at risk and a maximum temporal window of 50% including purely spatial clusters 2012-2022.

| Name of Sub County | Coordinates | Timeframe | Population | Observed case | Expected case | Annual cases/1000 | RR | LLR | P value |
| --- | --- | --- | --- | --- | --- | --- | --- | --- | --- |
| Dadaab and Fafi | (0.065322 N, 40.374216 E) / 90.14 km | 2012/1/1 to 2022/12/31 | 319292 | 4694 | 428.99 | 133.6 | 28.37 | 8679.669408 | < 0.001 |
| Nyatike, Suna West, Suba North, Uriri, Ndhiwa, Suna East and Awendo | (0.920871 S, 34.142828 E) / 44.64 km | 2015/1/1 to 2015/12/31 | 1029538 | 1296 | 125.66 | 126.0 | 12.30 | 1953.911976 | < 0.001 |
| Alego Usonga, Ugunja and Ugenya | (0.068122 N, 34.232069 E) / 19.41 km | 2016/1/1 to 2016/12/31 | 462938 | 554 | 56.66 | 119.4 | 10.49 | 783.164281 | < 0.001 |

**Figure 5:**
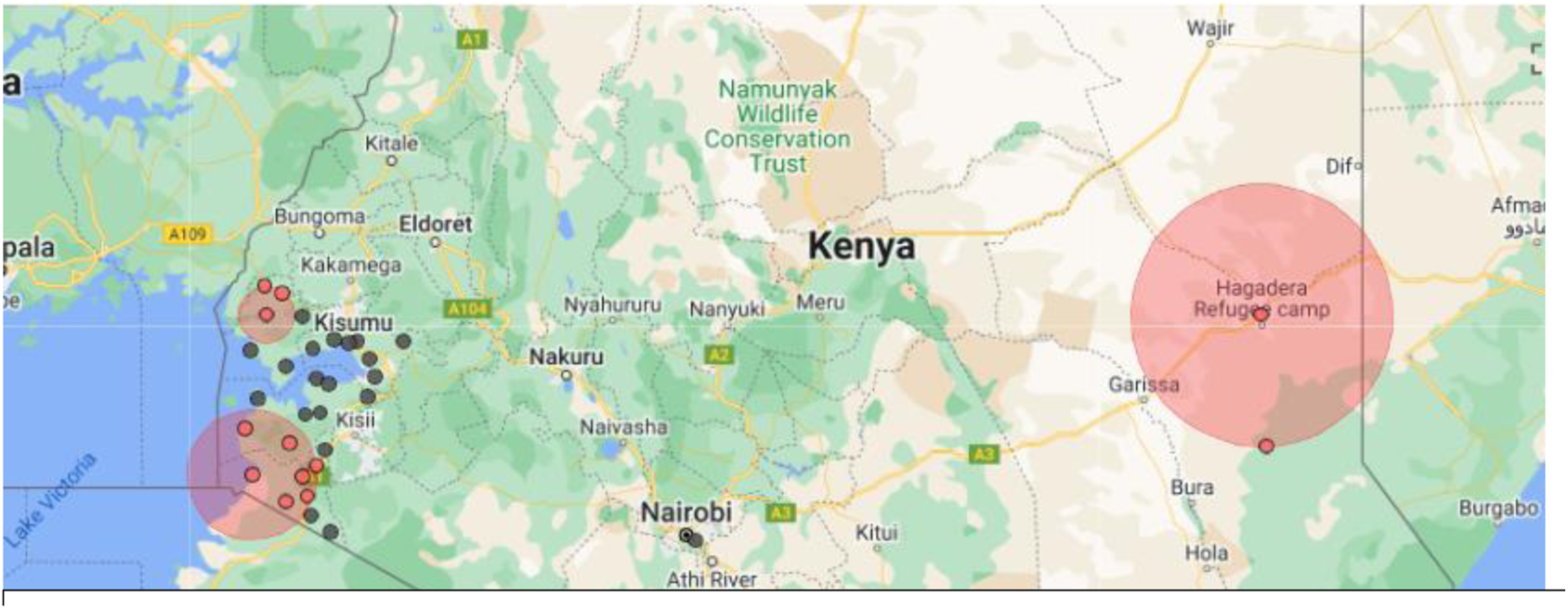
Map showing significant space-time clusters of cholera in selected high-risk areas.

The space-time analysis was adjusted to incorporate a circular spatial window with a radius of 50% of the population at risk and a maximum temporal window of 90%, including both purely spatial and purely temporal clusters. This modification allowed for a more comprehensive evaluation of the spatiotemporal patterns of cholera, considering a broader temporal window for cluster detection. As shown in Table 3, Dadaab and Fafi sub counties remained statistically significant high-risk clusters (p = 0.001) from 2015-2022.

**Table 3:** Significant high-risk spatial clusters of cholera in selected regions in Kenya, detected by retrospective space-time analysis using a circular spatial window with a radius of 50% of the population at risk and a maximum temporal window of 90% including purely spatial and purely temporal clusters, 2012-2022.

| Name of Subcounty | Coordinates | Timeframe | Population | Observed cases | Expected cases | Annual cases/1000 | RR | LLR | P value |
| --- | --- | --- | --- | --- | --- | --- | --- | --- | --- |
| Dadaab and Fafi | (0.065322 N, 40.374216 E) / 90.14 km | 2015/1/1 to 2022/12/31 | 319292 | 4603 | 311.97 | 180.2 | 37.62 | 9797.5157 | < 0.0001 |

### Spatial variation in temporal trends of cholera

There was a 0.654% overall annual increase in cholera cases with temporal and spatial variability (p<0.001) among the sub counties. Table 4 and Figure 6 below show Sub counties such as Dadaab and Fafi having an increasing trend within the window and a decreasing pattern outside the window.

**Table 4:** Spatial variation in temporal trends of cholera in the selected high-risk areas in Kenya, 2012-2022.

| Name of Subcounty | Observed cases | Expected cases | Relative risk | Annual cases/1000 | % cluster increase or decrease (Inside time trend) | % cluster increase or decrease (Outside time trend) | LLR |
| --- | --- | --- | --- | --- | --- | --- | --- |
| Dadaab, Fafi | 4694 | 428.99 | 28.37 | 133.6 | 10.850% annual increase | 15.439% annual decrease | 586.695196 |

**Figure 6:**
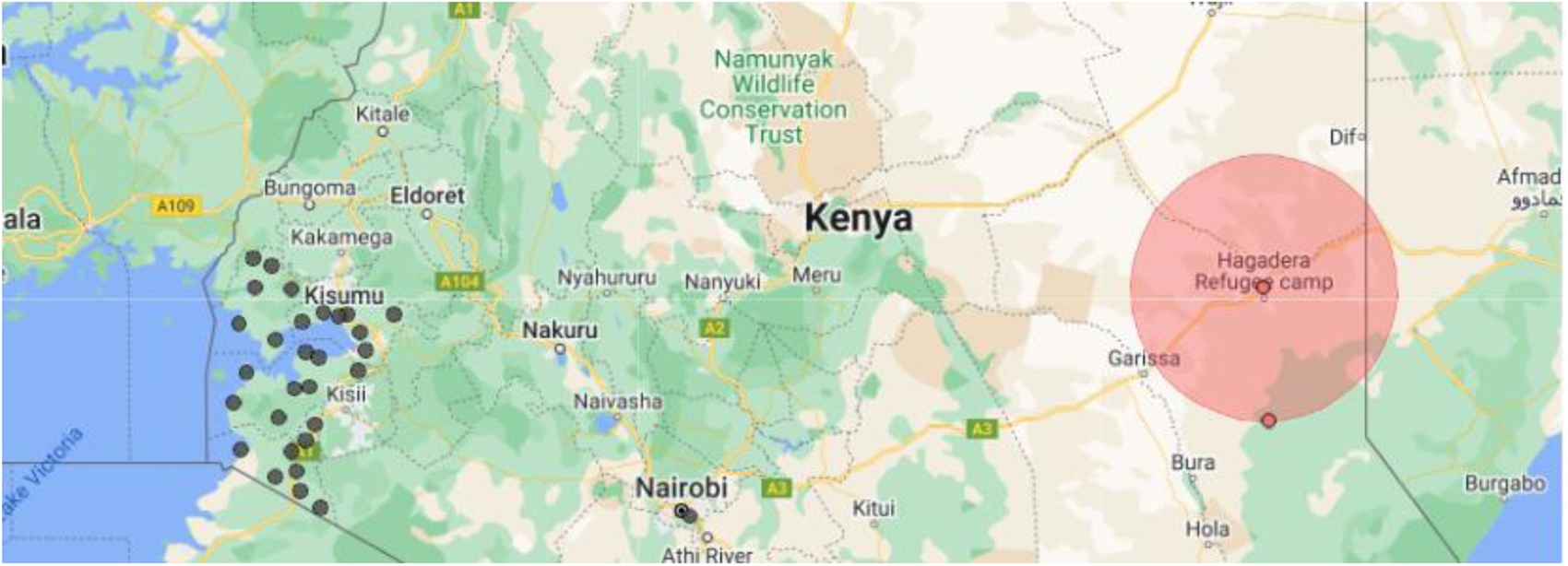
Map showing significant clusters of cholera in selected high-risk areas, Kenya, using a spatial variation in temporal trends analysis, 2012-2022.

## Discussion

During the study period, annual cholera epidemics followed a recurring pattern of waxing and waning, indicating a cyclicality in the occurrence of cholera outbreaks over time. In Kenya, these epidemic cycles have previously been reported to occur approximately every five to seven years lasting for two to three years (Kiama et al., 2023). In the current study, cholera cases waxed every three years in the selected high-risk areas. A similar trend has also been observed in countries such as India, Cameroon, and Bangladesh (Muzembo et al., 2022;Pascual et al., 2002;Ngwa et al., 2016). These countries have reported cholera epidemics that exhibit a recurring pattern with cycles ranging from 3 to 6 years. This similarity in cyclical occurrence of cholera outbreaks across multiple regions highlights a shared temporal pattern in the disease dynamics. By understanding these cycles, public health authorities can proactively plan interventions, allocate resources, and implement targeted measures to mitigate the impact of cholera outbreaks and work towards long-term prevention and control of the disease.

The peak of the cholera outbreak occurred in 2015, followed by a decline then a slight increase in number of cases in 2018. Additionally, there was another peak of cholera cases in 2022. The large number of cases observed in Kenya in 2015 was attributed to prolonged rainfall and subsequent flooding, which led to contamination of water sources and rapid spread of cholera (Moore et al., 2017). Conversely, the outbreak in 2022 was triggered by the impact of prolonged and severe drought, which resulted in water scarcity. Inadequate access to clean drinking water and poor sanitation exacerbated cholera transmission (WHO, 2023). In implementing appropriate prevention and control measures, it is important to consider such climatic factors that play a critical role in cholera outbreaks.

There is significant spatial and temporal variation in cholera patterns among the sub-counties studied. The observed differences in the spatial and temporal patterns of cholera can be attributed to the geographic variations and socio-economic factors within the study areas. Specifically, the study areas were characterized by diverse settings, including a refugee setting (Dadaab refugee camp), urban setting (informal settlements), and rural setting (counties bordering the Lake Victoria region). These different settings likely contribute to varying levels of cholera risk due to factors such as population density, access to clean household water and sanitation facilities, and healthcare resources. Understanding these geographic and socio-economic variations is crucial in tailoring effective cholera prevention and control strategies for each specific setting.

There were purely spatial and space-time statistically significant cholera clusters in selected high-risk areas in Kenya. Among these clusters, Dadaab and Fafi sub-counties stood out as the most significant, with a Relative Risk of 28.37 and a P-value <0.0001. This finding agrees with a previous study conducted on cholera hotspot mapping in Kenya using the GTFCC model, which also identified Dadaab as a high-risk sub-county (Kiama et al., 2023). Furthermore, similar findings have been reported in previous studies conducted in Ghana, Nigeria and Bangladesh, indicating the presence of possible spatial and temporal clustering of cholera (Ngwa et al., 2021; Debes et al., 2016;Osei & Duker, 2008). These consistent findings across different areas further strengthen the understanding of the epidemiology of cholera and its transmission dynamics, thus providing valuable insights for targeted prevention and control strategies.

During the study period, both the purely spatial analysis and the space-time analysis consistently identified Dadaab and Fafi sub-counties as having the highest number of observed cholera cases. When the space-time analysis was modified to include a maximum temporal window of 90% and incorporate temporal clusters, Dadaab and Fafi sub-counties continued to exhibit statistically significant purely spatial clusters from 2015 to 2022 (RR=37.62, P-value < 0.0001). This indicates a strong association between these sub counties and the occurrence of cholera cases, suggesting a persistent and significant cholera burden in these areas throughout the study period (see Table 3).

Dadaab refugee camp houses a significant number of refugees and asylum seekers, primarily originating from Somalia. Since its establishment in 1991, the average population of refugees in Dadaab has been steadily growing. The ongoing conflict, drought, and famine in Somalia continues to drive a substantial influx of people into the camp. Access to critical Water, Sanitation and Hygiene facilities therefore remain constrained as more people move into the already populated camps. This significantly increases the risk of cholera transmission among the refugee population. Drought can be identified as an additional contributing factor, as it has been linked to the extensive and prolonged transmission of cholera, often accompanied by high case fatality rates, particularly in arid and semi-arid regions. Previous studies have linked cholera outbreaks in Dadaab refugee camp to poor drainage systems and cross border movement (Golicha et al., 2018). This has also been observed in Zambia and Uganda where cholera cases have been linked to cross border movement (Bwire et al., 2017;Mwaba et al., 2020). A study by Breiman in 2009 additionally linked cholera outbreaks in Kakuma refugee camp in Kenya to storing drinking water at home in open containers and sharing latrines (Breiman et al., 2009). These findings stress on the importance of addressing these specific practices and implementing appropriate measures to prevent cholera outbreaks in refugee camps.

Nyakach, Nyando, Rachuonyo East, Kisumu East, Kisumu Central, Muhoroni, Rachuonyo North, Kisumu West, Suba South, Rangwe, Seme, Homabay Town, Rongo, Rarieda, Gem, and Awendo were identified as a cluster with a lower relative risk (RR=0.055), indicating a low rate of cholera cases in these areas. This spatial variation in cholera case distribution could be possibly explained by improved access to clean drinking water, proper sanitation and hygiene and adequate access to health facilities (Cowman et al., 2017).

In the years 2015 and 2016, Space-time analysis identified a cluster encompassing the areas surrounding Suba North, Uriri, Ndhiwa, Suna East, Alego Usonga, Ugenya, and Awendo. This cluster was not detected in the purely spatial analysis output, highlighting the significance of SaTScan in identifying recently emerging clusters. The use of space-time analysis allowed for the detection of dynamic patterns and trends that may not have been evident through purely spatial analysis alone. This finding emphasizes the importance of employing comprehensive analytical approaches, such as SaTScan, to capture the full extent of cluster patterns and improve our understanding of the temporal dynamics of cholera outbreaks.

This study reveals that cholera exhibits an overall increasing pattern of 0.654%, indicating a gradual rise in the number of cases over time. The increasing trend of cholera observed aligns with the findings reported by WHO in 2023 regarding the exponential rise in cholera cases in

Africa amidst a global surge. The WHO reports indicate that climate change has played a significant role in driving larger and more severe cholera outbreaks worldwide (WHO, 2023). The changing climate patterns, including increased temperatures and alterations in rainfall patterns, create favorable conditions for the growth and spread of *Vibrio cholerae*. There is therefore an urgent need for comprehensive strategies to address climate change impacts and mitigate risks associated with cholera outbreaks globally.

### Limitations of this study

Both suspected and confirmed cases were used in spatio-temporal analysis. This could result in overestimation of cholera cases. Laboratory confirmation of cases is crucial for accurate diagnosis and surveillance, but it may not always be feasible or accessible in resource-limited settings. Therefore, reported cholera cases may not provide a true picture of the disease burden in the selected high-risk areas.

Underestimation of cholera cases for individuals who did not seek treatment at public healthcare facilities was another limitation. Cholera cases that went unreported or were managed outside formal healthcare systems may not have been captured in the surveillance data. This could result in an incomplete representation of the true extent of cholera transmission within the population. Additionally, this study did not investigate potential risk factors contributing to cholera outbreaks. While the analysis focused on spatial and temporal patterns of cholera cases, other factors such as water and sanitation conditions, socio-economic factors, and hygiene practices could also play a role in the spread of the disease. Further research that incorporates investigation of these potential risk factors would provide a more comprehensive understanding of the determinants of cholera outbreaks in the high-risk areas.

## Conclusion

SaTScan analysis conducted in this study has provided valuable insights into the spatial and temporal patterns of cholera outbreaks in the high-risk areas in Kenya. The study identified statistically significant clusters of cholera cases, highlighting areas with elevated risk. The study also revealed the existence of both purely spatial and space-time clusters, emphasizing the need to consider both spatial and temporal dimensions in cholera surveillance and control efforts. This comprehensive approach enables the detection of emerging clusters and the implementation of timely interventions to prevent further spread of the disease.

Overall, this study contributes to the existing knowledge on cholera epidemiology and provides valuable insights to public health practitioners and policymakers in cholera prevention and control. It also serves as a basis for generating hypotheses for further research on contextual factors, including the impact of climate change on cholera occurrence and the risk factors related to water, hygiene, and sanitation in high-risk areas.

## Data Availability

All relevant data are within the manuscript and its supporting information files

## Notes

### Competing Interest Statement

The authors have declared no competing interest.

### Funding Statement

Yes

### Author Declarations

This research was approved by Kenyatta National Hospital_University of Nairobi Ethical Research Committee (KNH_UoN ERC) under approval number P731/09/2021

